# Impact of initial infected characteristics in an agent-based model of infectious respiratory disease: A methodological study

**DOI:** 10.1101/2025.07.02.25330763

**Authors:** Alexis Mandell, Mary Krauland, Mark Roberts

**Author notes:** **Corresponding Author**: Alexis Mandell, BA, Public Health Dynamics Laboratory, Department of Health Policy and Management, University of Pittsburgh School of Public Health Pittsburgh PA 15261.

## Abstract

There is limited existing literature about how infectious conditions are introduced in agent-based models (ABMs). This methodological study investigated the impact of the number, timing, and age of initial infected agents on the development of infectious disease outbreaks in ABMs, using influenza as an example. With an ABM, we modeled influenza in different size United States counties with different initial case characteristics including initial case number, timing, and age and calculated attack rate, season peak timing, and epidemic duration for each scenario. Increasing number of initial cases increased attack rate which plateaued at an initial case number proportional to population size. However, using a small initial case number resulted in many simulations with <1% attack rate. Introducing cases over time rather than at a single time point had minimal impact on attack rate but moved the season peak later in the season. Seeding infections in a younger age group increased the likelihood of a successful outbreak, attack rate, and epidemic duration.

In an ABM of an infectious respiratory disease, the characteristics of the initial infected cases impacted the resulting outbreak. The outbreak was accelerated, lengthened, or even nonexistent depending on how many, which, and when agents are first infected. Moving forward, it is important for ABM studies to justify seeding method and describe how it may impact results. Our study can be used to inform parameterization of initial conditions for ABMs of infectious diseases, enabling better forecasting of outbreaks and impacts of interventions.

## 1. Introduction

Infectious disease modeling is used widely for predicting influenza epidemics and analyzing current trends. Many types of models have been used historically to perform these tasks, and agent-based models (ABMs) have become increasingly common as computational power increases[1]. ABMs have a wide variety of individual, geospatial and global parameters and assumptions that can be adjusted to change the characteristics of a modeled disease. Because of the large number of parameters, estimating how each of those impacts a simulation is complex. In an infectious disease model, an outbreak does not start naturally in the simulation population but must be externally “introduced” or “seeded”. As such some of the many modeling parameters control how the infectious condition is introduced into the population to start disease transmission. In a perfect world, when modeling influenza, the modeler would know the exact timing and number of cases brought into a population by travel or other means at the beginning of the season, but this is rarely the case. However, ABMs have the advantage of granular, individual-level detail that allows for initial insertion of cases (referred to here as seeding) to mimic how it is hypothesized to occur in real life.

To our knowledge, there have been no formal investigations of the effect of the initial seeding parameters on the results generated by an ABM. Several studies have discussed how seeding initial cases impacts the output of metapopulation network models of infectious conditions[2,3,4,5,6]. Conversations about seeding an ABM in the literature are often limited to a few sentences in the methods section with little explanation of rationale[7,8,9,10,11,12]. A few identified studies have more detailed discussions of seeding and focus mainly on location of initial cases. In an ABM built for the Australian population, Cliff et.al. seed at international airports at levels proportional to air traffic flow due to Australia’s geographic isolation from disease spread in the rest of the world[13]. Ajelli et.al. vary the seeding municipality as part of their analysis of adding unstructured contacts to an ABM[14]. Moving forward, it is important to understand how different parameters for initial infections in an ABM impact the results so our models best reflect biological reality of disease dynamics.

The purpose of this paper is to investigate the impact of the number, timing and age of initial cases on the development of influenza epidemics in an ABM. We used the Framework for Reconstructing Epidemiological Dynamics (FRED) platform to study the effect of changing initial case characteristics across different values of basic reproduction number in an influenza model that models transmission through mixing groups.

## 2. Methods

The Framework for Reconstructing Epidemiological Dynamics (FRED) is an ABM platform that uses a synthetic population based on the 2010 United States Census. Each FRED model is created with one or more infectious conditions that spread through interactions between agents in their homes, schools, workplaces, and neighborhoods. Each condition includes a set of states and rules for transitions between those states. FRED has previously been described in detail[15,16,17,18].

This study used a modified SEIR influenza model with added states for presymptomatic infections, asymptomatic infections, hospitalization, and death. Infected agents are hospitalized and die at rates obtained from published Centers for Disease Control and Prevention (CDC) data[19,20]. Agents are vaccinated at the national coverage rate published by the CDC for the 2019-20 influenza season, and vaccination is specified to be 40% effective at preventing infection. 25% of influenza cases in the model are asymptomatic[21,22]. To accommodate the seasonal forcing observed in influenza, models include a seasonal transmissibility parameter that increases or decreases transmissibility depending on how far the current day is from the winter solstice[23]. This is intended to replicate the identified seasonality of respiratory diseases in the United States. We introduce influenza into the population starting on October 15th. Further model details are provided in the Supplementary Materials.

It is important to note that infectivity, typically measured for an infectious disease by R_0_ (or R_eff_ in the case of existing immunity) is not an input in FRED, which uses a combination of the transmissibility of the virus, the length of contact, and susceptibility of the recipient to determine if an infection takes place. Therefore, R_0_ and R_eff_ are *results* of our model, and the transmissibility parameter can be calibrated to produce a given R_0_ or R_eff_. We used literature sources to choose the goal R values for this simulation. A systematic review of R estimates for influenza found a range from 1.27 for seasonal influenza to 1.80 or higher for pandemic influenza strains such as that from the 1918 influenza pandemic[24]. We chose transmissibility parameters that produce the desired R_0_ for the typical range of seasonal influenza. In this analysis, we present results using parameters appropriate for H1N1 and H3N2 seasonal influenza. The R_0_ estimates and a sensitivity analysis with other transmissibility values along with further explanation can be found in the Table S2.

The chosen parameters for seed size, age, and timing are described in Table 1. Seed size refers to the number of initial cases inserted in the population. Each seeding scenario was simulated 100 times in FRED to account for stochastic variation. Attack rate, the epidemic duration, and the day of maximum incidence were calculated as an average across runs and for each simulation. Attack rate (AR) was calculated as the total number of infections over the total population. Epidemic duration was calculated as the number of days with greater than 5 incident influenza cases per 100,000 population, a threshold chosen to represent above off-season rates of influenza. Failed outbreak simulations were also recorded to evaluate how seeding characteristics impact their incidence. A failed outbreak was defined as a simulation in which the AR is less than 1%. FRED reports both asymptomatic and symptomatic cases, so symptomatic attack rates are 25% lower than the reported overall attack rate.

**Table 1:** Model parameters for each of the seeding characteristics. Each model was run in four counties with transmissibility values appropriate for H1N1 influenza and for H3N2 influenza. Influenza was introduced to the population starting on October 15^th^ and vaccination began on September 15^th^ in all models.

| Seeding Characteristic | Modeled Scenarios |
| --- | --- |
| Number of cases (seed size) | 1-100 initial cases by increments of 10 |
| Age | 50 initial cases in school-age (5-17) group<br>50 initial cases in working-age (18-64) group<br>50 initial cases in older age (50+) group |
| Timing | 50 initial cases seeded on the same day<br>One case seeded per day for 50 days<br>5 cases seeded each week for 10 weeks |

Four counties in Pennsylvania were chosen to create a mix of population sizes and rural, suburban, and urban populations. Philadelphia County is an urban county with a population of 1.5 million individuals. Dauphin County is suburban and has a population of around 300,000 individuals. Jefferson County is a rural county with around 45,000 individuals. Allegheny County has a population of 1.2 million individuals and contains a mix of urban and suburban areas. Additionally, Allegheny County is a good representation of the United States as a whole in terms of demographics and age structure. More details on the characteristics of the modeled counties can be found in the Table S3 and S4.

## 3. Results

### 3.1 Number of Initial Cases

As seed size increases, attack rate increases but plateaus at a seed size that is proportional to the county population size (Figure 1). The attack rate plateaus more quickly in smaller, less dense counties than in large urban counties. In larger counties, attack rates plateau at around 50 seed cases, and in smaller counties at 20-30. Increasing the seed size had a smaller effect as the transmissibility of the influenza strain increased.

**Figure 1:**
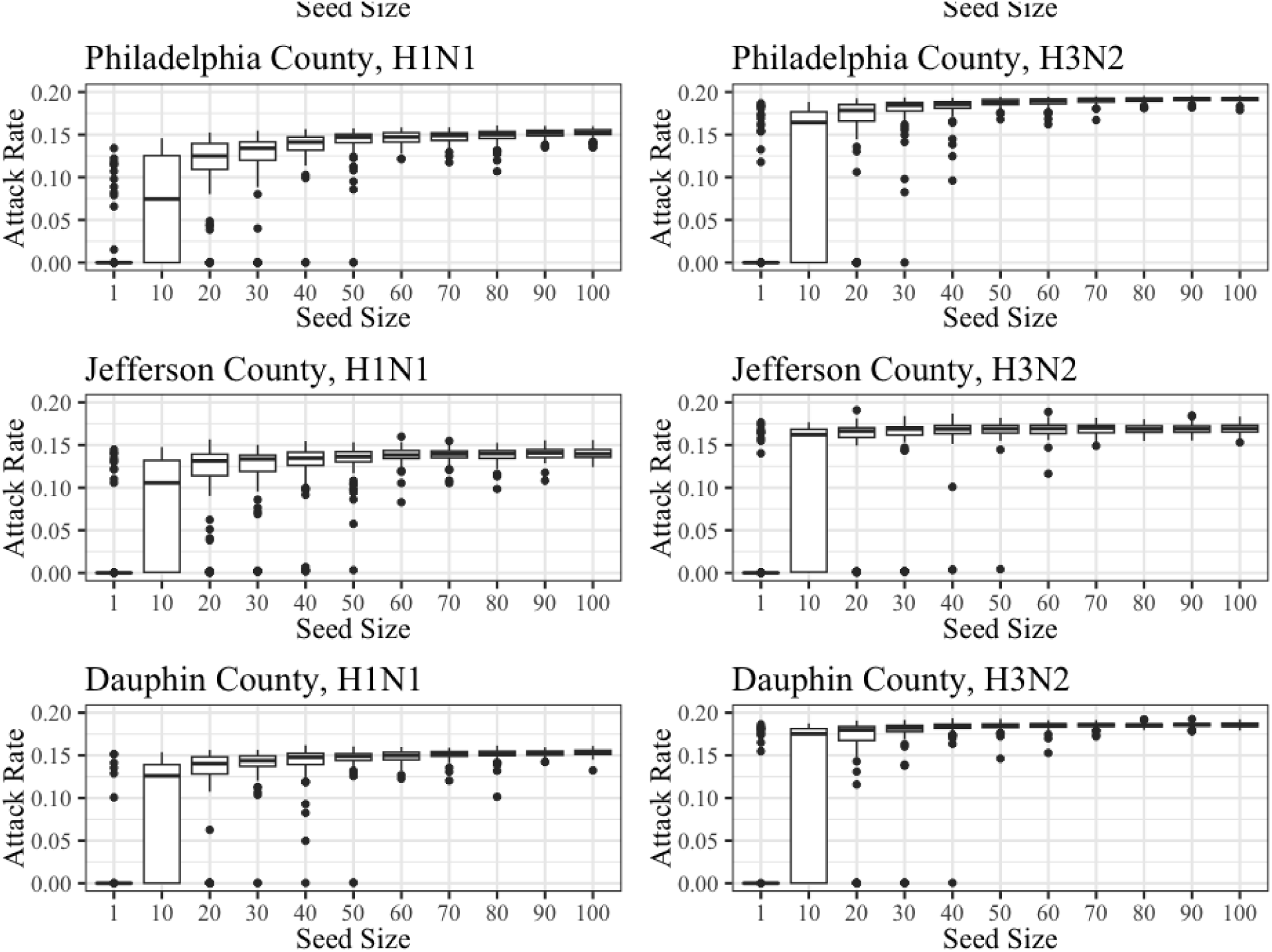
Distribution of overall attack rate (asymptomatic and symptomatic cases) over 100 simulations for each seed size value and influenza subtype in Allegheny, Philadelphia, Jefferson, and Dauphin counties in Pennsylvania. Seed size refers to the number of infected cases inserted into the population at the start of the simulation.

The data in Figure 1 includes simulations with failed outbreaks (AR < 1%). Because transmission is stochastic, some introductions, especially in small counties, will not produce an outbreak. Including simulations with outbreaks below the threshold for success lowered the mean AR across 100 simulations for a given seed size. When those were excluded, the mean AR increased between 6 and 15% (Figure S3). The increase was larger for the lower seed sizes because more simulations with lower seed size resulted in failed outbreaks. For example, for an H1N1-like strain in Allegheny County, 91% of simulations with a seed size of one resulted in a failed outbreak; for the same strain in Jefferson County, 90% of such simulations resulted in a failed outbreak. For seed sizes greater than 40 in every county, every simulation resulted in an outbreak. When excluding simulations below the successful outbreak threshold, AR still increased with seed size, but the effect was attenuated. Exclusion of simulations with failed outbreaks decreases the standard deviation of the AR.

The day of maximum incidence moved earlier in the season as the seed size increased for both the small and large populations. The smaller counties have an earlier peak than the larger counties. Similar to the mean AR measure, at a seed size of 50 or 60 initial cases in the larger counties, the change in mean peak date with increasing seed size decreased, and this effect was more pronounced for the more transmissible subtype (H3N2). Epidemic duration as measured was shortest for a seed size of 1 case. It increased by about two months for a seed size of 10 compared to 1 and about 1-2 months for a seed size of 20 compared to 10. Epidemic duration plateaued at a seed size of around 50 cases (Table S5).

### 3.2 Initial Case Age

Across all four counties, attack rates ranged from 19.0% to 28.2% for H3N2-like influenza and from 14.5% to 25.1% for H1N1-like influenza (Figure 2). Seeding in the 50+ age group always resulted in the lowest attack rate, on average 13.6% smaller than the under 18 seed runs for H3N2 and 23.0% smaller than the under 18 seed runs for H1N1. Seeding in the under 18 age group versus in the 18-49 age group created a smaller difference in attack rate, on average 2.4% difference for H3N2 and 4.0% difference for H1N1. As part of transmission by mixing group, FRED has place-based contact rates that are higher for classrooms, schools, offices, and workplaces then in the neighborhood or at home. Because of this, agents that spend time in schools or at work have more contacts than agents that do not which likely contributes to the larger attack rates when seeding in the school-age or working-age population.

**Figure 2:**
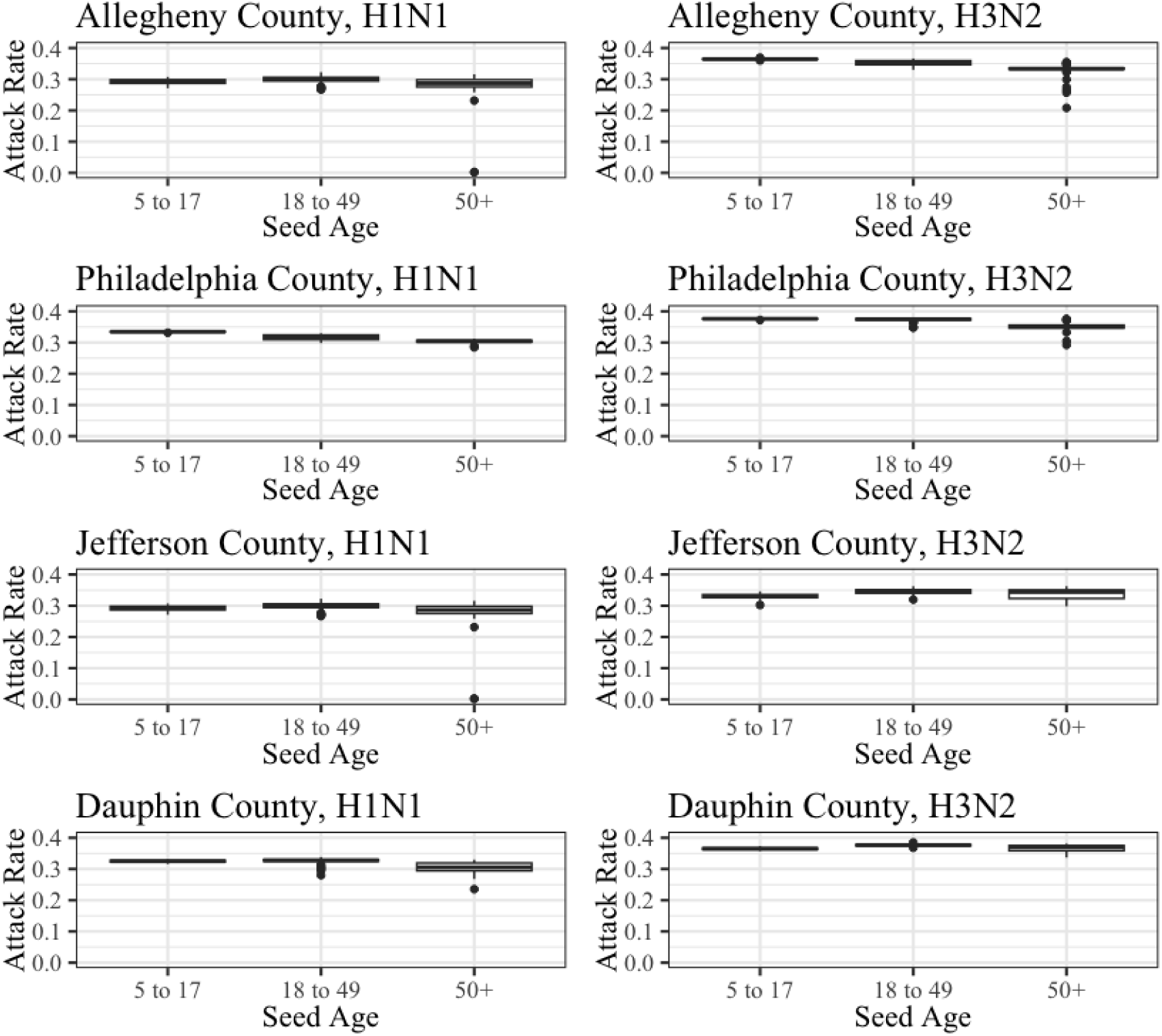
Distribution of overall attack rate (asymptomatic and symptomatic cases) over 100 simulations for each seed age group and influenza subtype in Allegheny, Philadelphia, Jefferson, and Dauphin counties in Pennsylvania.

The day of maximum incidence moved later in the season as the age of the seed agents increased across all four counties and for both influenza subtypes. The under 18 seed simulations peaked about two weeks earlier than the 18-49 seed simulations which peaked about one week earlier than the 50+ seed simulations. Epidemic duration was the shortest with seed cases in the 50+ age group (Table S6). The season was between 1 and 3 weeks shorter when seeded with agents in the 50+ age group compared to younger seed agents. The longest duration varied between the under 18 seed and the 18 to 49 seed scenarios. The difference in season length between these two groups was less than one week.

### 3.3 Initial Case Timing

The method of seed timing had minimal impact on the attack rate (Table 2). Across all of the counties, the single seed resulted in a higher burden of infections than either daily or weekly seeding, but the difference between strategies was smaller for the higher transmissibility virus. The percent difference in total number of cases between strategies ranged from 0.5% to 13%.

**Table 2:** Overall attack rates (asymptomatic and symptomatic) for each of the seed timing simulations for both the H3N2- and H1N1-like strains.

| County | Single Seed | Daily Seed | Weekly Seed |
| --- | --- | --- | --- |
| Allegheny | H3N2-like strain: 19.1%<br>H1N1-like strain: 12.4% | H3N2-like strain: 17.9%<br>H1N1-like strain: 10.8% | H3N2-like strain: 17.8%<br>H1N1-like strain: 11.3% |
| Dauphin | H3N2-like strain: 18.6%<br>H1N1-like strain: 14.9% | H3N2-like strain: 18.4%<br>H1N1-like strain: 14.7% | H3N2-like strain: 18.6%<br>H1N1-like strain: 13.9% |
| Jefferson | H3N2-like strain: 17.9%<br>H1N1-like strain: 12.0% | H3N2-like strain: 17.5%<br>H1N1-like strain: 11.8% | H3N2-like strain: 17.1%<br>H1N1-like strain: 12.1% |
| Philadelphia | H3N2-like strain: 18.9%<br>H1N1-like strain: 14.5% | H3N2-like strain: 18.1%<br>H1N1-like strain: 13.2% | H3N2-like strain: 18.0%<br>H1N1-like strain: 13.3% |

Seed timing had minimal impact on the epidemic duration as we defined it. For both counties the number of days with more than 5 cases of influenza per 100,000 differed by less than 1 week between any of the seed timing strategies (Table S7). Multiple seeds over time rather than a single seed moved the day of maximum incidence later in the season. A single seed resulted in the earliest peak followed by daily seeding, and weekly seeding resulted in the latest peak.

Seeding over the course of weeks spread infections over a longer period of time, resulting in lower burden on the day of maximum incidence (Figure 3).

**Figure 3:**
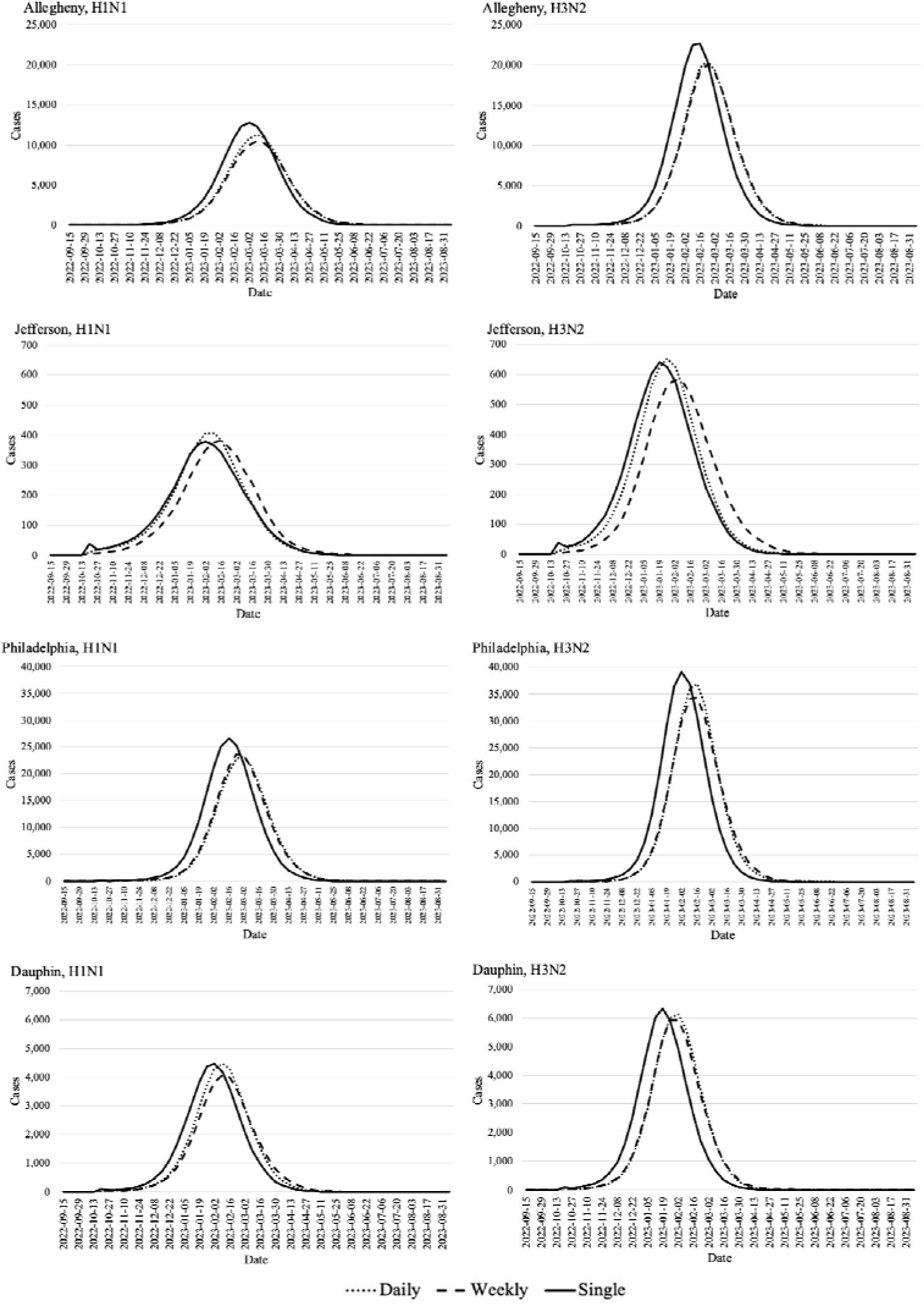
Allegheny, Jefferson, Philadelphia, and Dauphin County, PA epidemic curves for each seed timing strategy and influenza A subtype. Epidemic curves are from mean weekly incident cases from 100 simulations in FRED. Seed timing strategies were a single import of seed cases on one day, weekly imports of seed cases, and daily imports of seed cases.

## 4. Discussion

This study’s purpose is to investigate the effects of seeding characteristics on the performance of ABMs of infectious disease, using influenza as an example. Our results suggest that no matter the timing of the initial seed, eventually agent-to-agent transmission overtakes the initial insertion of cases and results in similar overall attack rates. However, using a very small number of initial cases (<20), especially in a large population, can result in a large number of simulation scenarios that produce no outbreak, which may impact results. The age of initial cases had a larger impact on the resulting attack rate, but this effect can be controlled for by seeding randomly and simulating the outbreak many times to avoid many seed cases in any single age group. If simulating an infectious disease outbreak that begins in children or the elderly, it is worth considering how seed age impacts results independent of other factors when calibrating the model and whether it is consistent with reality.

Moving forward, it is important to specify in ABM studies that analyze multiple runs of the same model how an outbreak was seeded, whether simulations with failed outbreaks occurred, and whether or not they were excluded from analysis. In our study, the characteristics of the initial cases had a larger impact on the duration and timing of the resulting outbreak than on the size of the outbreak. The day of maximum incidence moves with seed size, age, and timing. When designing ABM models to forecast peak dates or when modeling time-specific policies that depend on a threshold level of infections it is important to choose seeding strategies that replicate reality and do not introduce artifacts that negatively impact the robustness of results.

Methodological studies of ABM are important to assist in creating realistic representations of disease. If more detailed data was collected about the first detections of influenza each season, it would assist in developing more accurate models. ABMs in particular would benefit because of their ability to incorporate detailed data into their parameters. Understanding and controlling each parameter, including seeding characteristics like size, timing, and age, advances this goal and allows for better modeling of interventions. By modeling with different initial case characteristics, we may gain insights for influenza surveillance, such as likely outbreak trajectories based on the age of the people first infected or how many infections are necessary to start widespread transmission. Because the introduction of an infectious condition in a model is by nature artificial, conducting these investigations also helps to create models that better reflect natural progressions of outbreaks.

## 5. Study Limitations

This study has several limitations. Models are always an oversimplification of what happens in real life. Influenza is difficult to predict and highly variable from year to year. The models used in this study and their parameters came from the current best available data, but vaccine effectiveness and influenza burden vary each season. In addition, surveillance for influenza in the United States is not all encompassing, making estimates of vaccine effectiveness and attack rate difficult. Again, we used best available data from the CDC when modeling, but there could be discrepancies between published data and actual values. While influenza is variable across seasons, that is unlikely to decrease the validity of this investigation.

While this study focuses specifically on influenza in an ABM that does transmission by mixing group, the findings may be generalizable to modeling of other diseases with a similar mode of transmission. They could also be more broadly applicable to other modeling methods, but further research needs to be done on the impact of varying seed characteristics in other types of models. Conversely, some of the findings presented here could be artifacts of the FRED modeling system or specific model implementations in the influenza model and not generalizable to other model types or systems.

## Data Availability

All data produced in the present study are available upon reasonable request to the authors.

## Acknowledgements

This work was supported by the Center for Disease Control and Prevention U01-IP001141-01. The contents are those of the authors and do not necessarily represent the official views of, nor an endorsement, by CDC/HHS, or the U.S. Government.

