## Supplementary Materials for "Impact of initial infected characteristics in an agent-based model of infectious respiratory disease: A methodological study"

Table of Contents

1. FRED Influenza Model Details
2. Basic Reproduction Number Estimates for the FRED Transmissibility Parameter
3. Attack Rate Results Excluding Simulations with No Outbreak
4. Results of sensitivity analysis for varying initial case numbers across other FRED transmissibility values
5. Characteristics of Included Counties
6. Epidemic Duration Results
7. References

1. *FRED Influenza Model Details*

**Figure S2**: A state diagram for the FRED influenza model used in this study

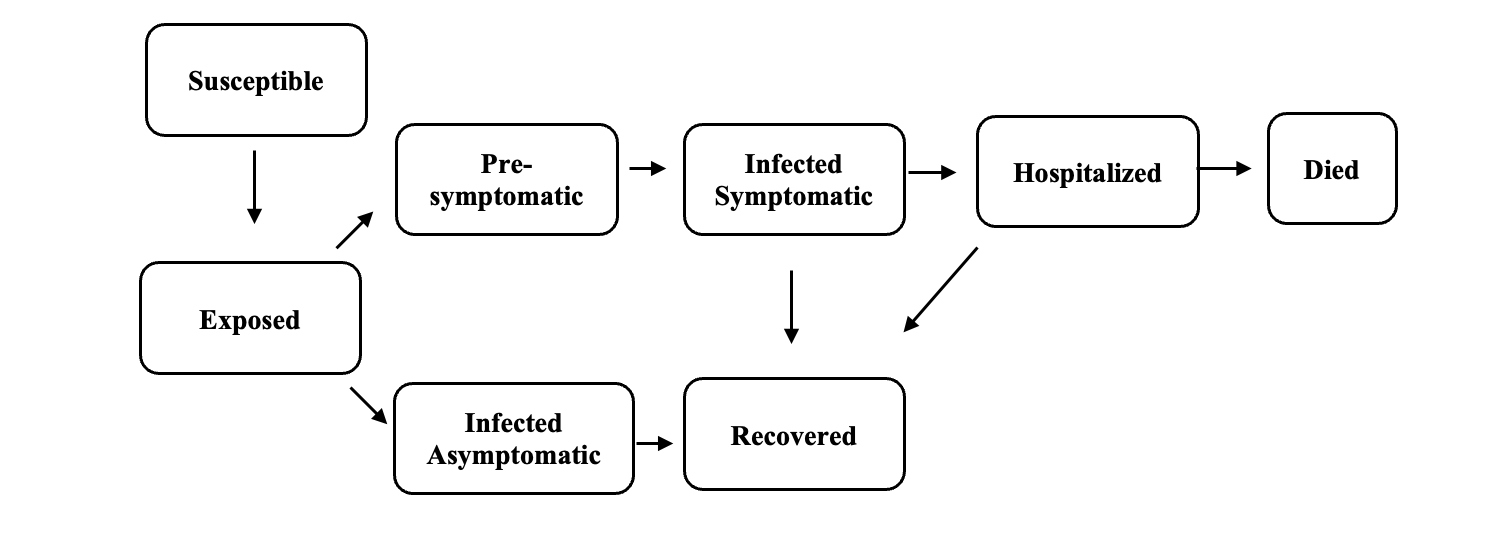

**Figure S1**: A timeline of events in the FRED influenza model used for this study

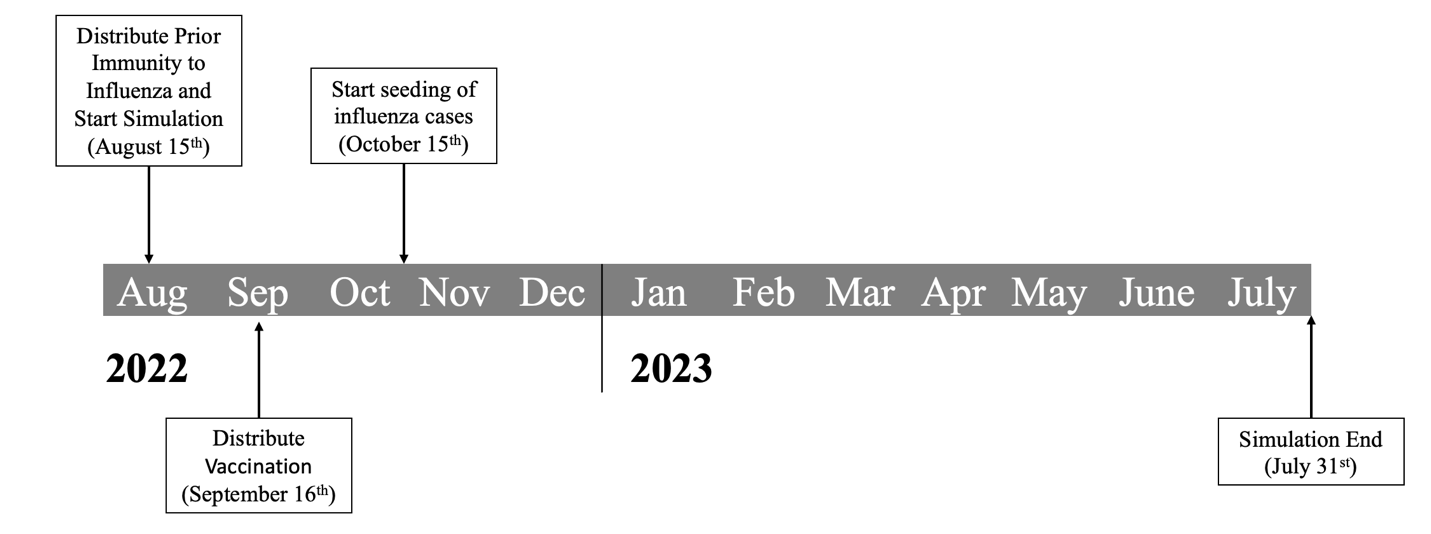

The FRED influenza model is a modified SEIR model with added states for asymptomatic infections, hospitalization, and death. Asymptomatic agents receive a 50% reduction in their transmissibility. Once infected, an agent’s susceptibility to future infection decreases to 0, and this protection wanes at a rate of 3% per month. Symptomatic infected agents are hospitalized and die at a rate based on their age group published by the Centers for Disease Control and Prevention (CDC). (1, 2)

Agents are vaccinated within a period of six weeks starting on September 16^th^ at rates based on age group published by the CDC for the 2019-20 influenza season. Two weeks after vaccination, agents receive 40% protection from infection. Vaccine protection wanes at a rate of 7% per month until the end of the simulation.

| **Table S1**: Vaccination coverage for agents based on CDC influenza vaccination coverage rates for the 2019-20 influenza season (3) | |
| --- | --- |
| CDC Age Group (years) | Vaccination Coverage (2019-20 season) |
| 0.5-17 | 0.638 |
| 18-49 | 0.384 |
| 50-64 | 0.506 |
| 65+ | 0.698 |

2. *Basic Reproduction Number Estimates for the FRED Transmissibility Parameter*

Infectivity, typically measured for an infectious disease by R_0_ (or R_eff_ in the case of existing immunity) is not an input in FRED, which uses a combination of the transmissibility of the virus, the length of contact, and susceptibility of the recipient to determine if an infection takes place. Therefore, R_0_ and R_eff_ are results of our model, and the transmissibility parameter can be calibrated to produce a given R_0_ or R_eff_.

To produce the estimations of the implied basic reproduction numbers in the table below, we ran our influenza model without vaccination or prior immunity in Allegheny County, Pennsylvania to produce an R_0_ estimate for each transmissibility value (Table S2). For the results shown in this study, the R_eff_ for each of these transmissibility values would be lower due to the inclusion of vaccination and prior immunity for influenza. When modeling H1N1 subtype, we used a transmissibility value of 0.65, and when modeling H3N2 subtype, we used a transmissibility value of 0.70.

| **Table S2**: FRED transmissibility parameter and estimates of the implied R_0_ from 100 simulations without vaccination or prior immunity in Allegheny County, Pennsylvania | |
| --- | --- |
| FRED Transmissibility Parameter | Implied Basic Reproduction Number (R_0_) |
| 0.60 | 1.08 |
| 0.65 | 1.32 |
| 0.70 | 1.48 |
| 0.75 | 1.68 |
| 0.80 | 1.74 |

3. *Attack Rate Results Excluding Simulations with No Outbreak*

Occasionally, a FRED influenza simulation can result in no transmission of cases beyond the initial import. This is more common when using a small seed size (<30 cases). Figure S3 shows the difference in resulting attack rate across seed size excluding failed simulations. For a seed size of 1, up to 91% of the simulations resulted in no outbreak.

**Figure S3**: Distribution of AR for each seed size when excluding simulations with a failed outbreak.

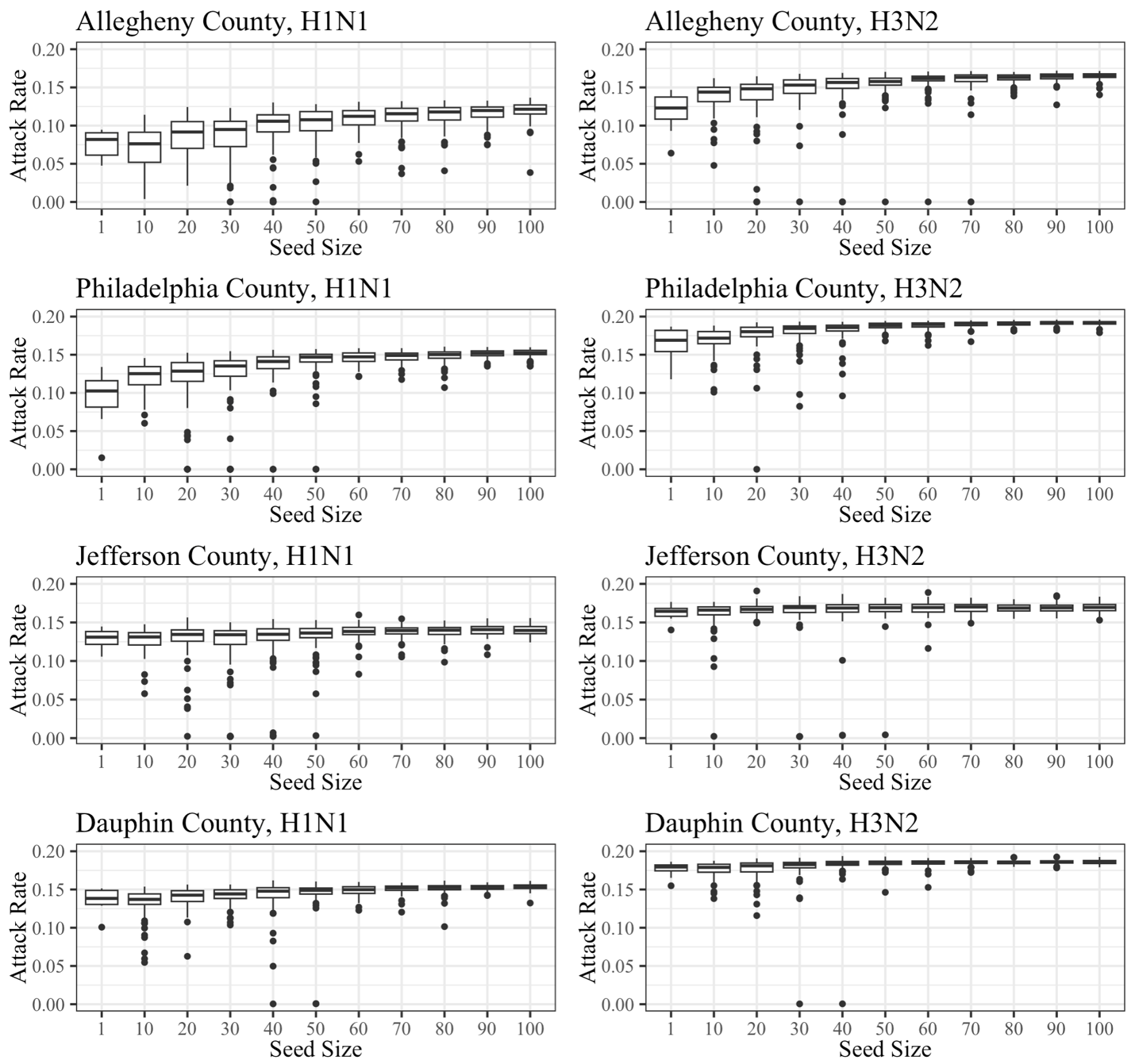

4. *Results of sensitivity analysis for varying initial case numbers across other ~R_0_*

For a limited sensitivity analysis for the transmissibility parameter, we model with smaller and larger approximate R_0_ values. compared to the main analysis for each seed size in Allegheny, Dauphin, Jefferson, and Philadelphia County. Each county, ~R_0_, and seed size combination was modeled 100 times in FRED. As the ~R_0_ increases, the attack rates increases and the standard deviation of attack rate decreases.

**Figure S4**: Distribution of attack rates from ~R_0_=1.08 across seed sizes and counties

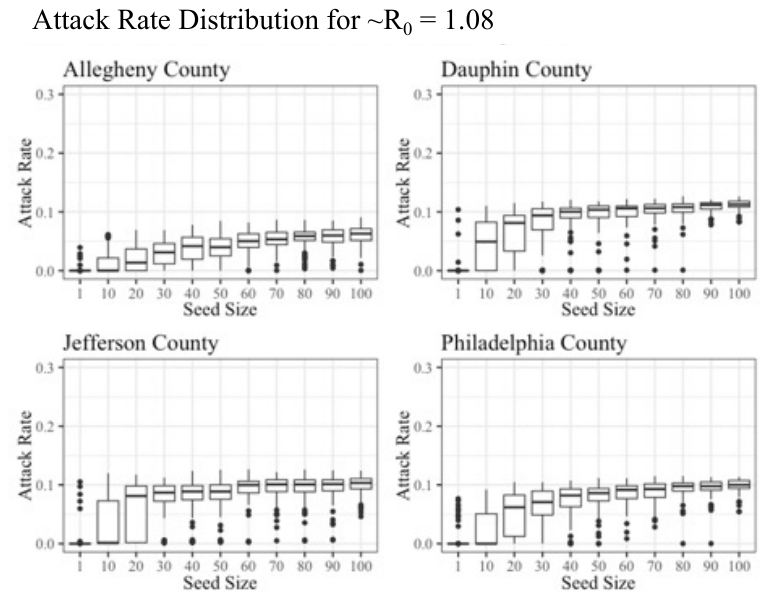

**Figure S5**: Distribution of attack rates from ~R_0_=1.68 across seed sizes and counties

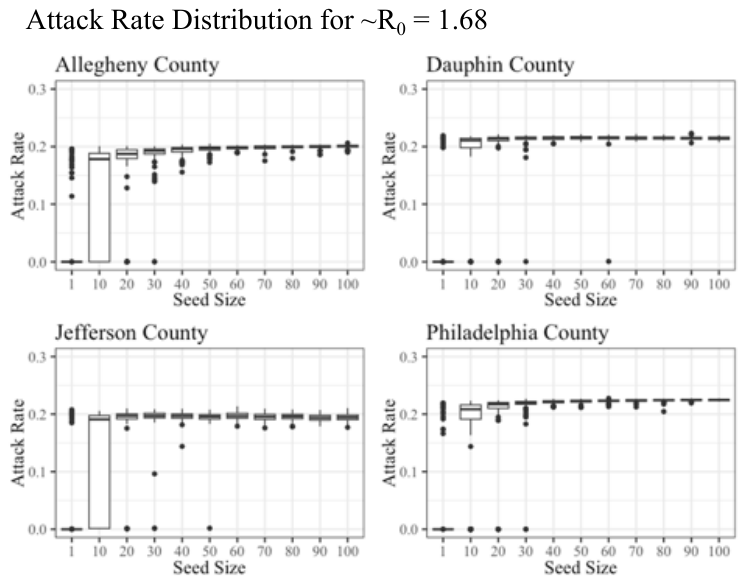

**Figure S6**: Distribution of attack rates from ~R_0_=1.74 across seed sizes and counties

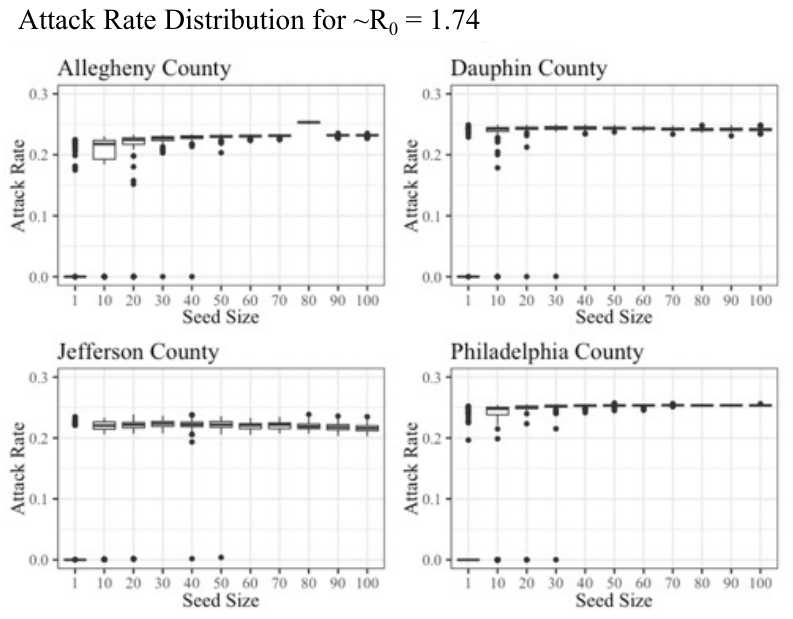

5. *Characteristics of Included Counties*

The four counties included in the analysis were chosen to include a range of county size and population density (Table S3). Also, Allegheny County has a comparable age distribution to the United States as a whole (Table S4).

| **Table S3**: Overall population, population density, and median age for the four counties included in the analysis. | | | |
| --- | --- | --- | --- |
| County Name | Population | Pop. Density (people/sq. mile) | Median Age |
| Allegheny | 1,218,695 | 1700 | 41 |
| Dauphin | 289,234 | 479 | 40 |
| Jefferson | 45,318 | 68 | 44 |
| Philadelphia | 1,508,447 | 4,456 | 35 |

| **Table S4**: Age group breakdown of the Allegheny County, PA FRED population and the United States 2020 population. | | | | |
| --- | --- | --- | --- | --- |
| Age Group | FRED Allegheny County Synthetic Population Age Makeup | Number of Agents in Age Group in FRED Allegheny County Population | United States Population Age Makeup (2020) | Number of People in Age Group in the United States Population (2020) |
| 0 to 4 | 5.17% | 63,016 | 4.39% | 14,550,623 |
| 5 to 17 | 15.30% | 186,501 | 16.23% | 53,794,218 |
| 18 to 49 | 41.49% | 505,676 | 41.91% | 138,910,394 |
| 50 to 64 | 21.05% | 256,540 | 18.68% | 61,914,726 |
| 65+ | 16.98% | 206,962 | 17.70% | 58,666,523 |

6. *Epidemic Duration Results*

For this study, epidemic duration was defined as the number of days with greater than 5 incident influenza cases per 100,000 population. We chose this threshold to represent greater than off-season rates of influenza. Tables S5-S7 contain the epidemic duration results for each of the seeding characteristic simulations.

| **Table S5**: Mean epidemic duration and standard deviation (days) across 100 runs for each seed size simulation | | | | | |
| --- | --- | --- | --- | --- | --- |
| Influenza Subtype (~R_0_) | Seed Size | Allegheny County | Philadelphia County | Dauphin County | Jefferson County |
| H1N1 (1.32) | 1 | 13 (41) | 16 (43) | 9 (34) | 16 (46) |
|  | 10 | 71 (69) | 70 (67) | 90 (67) | 88 (79) |
|  | 20 | 106 (60) | 120 (42) | 125 (46) | 135 (61) |
|  | 30 | 126 (43) | 123 (39) | 141 (21) | 150 (48) |
|  | 40 | 131 (39) | 133 (19) | 145 (16) | 157 (37) |
|  | 50 | 141 (15) | 134 (19) | 146 (22) | 165 (10) |
|  | 60 | 143 (4) | 137 (4) | 150 (7) | 166 (11) |
|  | 70 | 145 (4) | 137 (3) | 152 (7) | 165 (12) |
|  | 80 | 144 (4) | 138 (4) | 156 (7) | 163 (11) |
|  | 90 | 146 (4) | 139 (4) | 157 (6) | 160 (11) |
|  | 100 | 146 (4) | 139 (4) | 160 (5) | 160 (9) |
| H3N2 (1.48) | 1 | 17 (45) | 20 (46) | 14 (41) | 14 (42) |
|  | 10 | 91 (64) | 85 (61) | 97 (59) | 107 (69) |
|  | 20 | 113 (50) | 111 (45) | 122 (41) | 135 (47) |
|  | 30 | 127 (35) | 128 (14) | 131 (30) | 145 (38) |
|  | 40 | 133 (19) | 129 (4) | 139 (15) | 152 (21) |
|  | 50 | 135 (14) | 130 (3) | 142 (6) | 152 (16) |
|  | 60 | 136 (14) | 131 (3) | 144 (6) | 153 (11) |
|  | 70 | 137 (14) | 131 (3) | 146 (6) | 152 (10) |
|  | 80 | 138 (4) | 133 (4) | 148 (5) | 151 (10) |
|  | 90 | 139 (3) | 134 (4) | 149 (5) | 149 (10) |
|  | 100 | 140 (4) | 134 (3) | 151 (5) | 147 (8) |

| **Table S6**: Mean epidemic duration and standard deviation (days) across 100 runs for each seed age group simulation | | | | | |
| --- | --- | --- | --- | --- | --- |
| Influenza Subtype (~R_0_*) | Seed Age Group | Allegheny County | Philadelphia County | Dauphin County | Jefferson County |
| H1N1 (1.32) | Under 18 | 138 (5) | 130 (4) | 143 (5) | 139 (7) |
|  | 18 to 49 | 145 (4) | 133 (5) | 138 (5) | 142 (29) |
|  | 50 and older | 128 (48) | 128 (36) | 118 (50) | 102 (68) |
| H3N2 (1.48) | Under 18 | 139 (4) | 130 (4) | 144 (5) | 140 (6) |
|  | 18 to 49 | 138 (4) | 127 (2) | 139 (4) | 145 (22) |
|  | 50 and older | 137 (25) | 123 (34) | 127 (32) | 112 (62) |

*~R_0_ refers to the estimated basic reproduction number which was obtained by running a model with a given transmissibility value and no vaccination or prior immunity

| **Table S7**: Mean epidemic duration and standard deviation (days) across 100 runs for each seed timing simulation | | | | | |
| --- | --- | --- | --- | --- | --- |
| Influenza Subtype (~R_0_*) | Seed Timing | Allegheny County | Philadelphia County | Dauphin County | Jefferson County |
| H1N1 (1.32) | Single | 137 (25) | 135 (3) | 147 (7) | 131 (66) |
|  | Daily | 136 (22) | 132 (3) | 140 (6) | 127 (65) |
|  | Weekly | 136 (20) | 126 (4) | 137 (15) | 123 (55) |
| H3N2 (1.48) | Single | 135 (3) | 127 (13) | 140 (15) | 134 (49) |
|  | Daily | 133 (4) | 127 (5) | 132 (5) | 130 (51) |
|  | Weekly | 133 (4) | 126 (4) | 133 (7) | 129 (43) |

*~R_0_ refers to the estimated basic reproduction number which was obtained by running a model with a given transmissibility value and no vaccination or prior immunity

*7. References*

1. FluSurv-NET: Influenza Hospitalization Surveillance Network, Centers for Disease Control and Prevention. <https://gis.cdc.gov/GRASP/Fluview/FluHospRates.html>. Accessed on 24 August, 2023.

2. Centers for Disease Control and Prevention. “Estimated Flu-Related Illnesses, Medical visits, Hospitalizations, and Deaths in the United States – 2019-2020 Flu Season.” <https://www.cdc.gov/flu/about/burden/2019-2020.html>. Accessed on 24 August 2023.

3.Flu Vax View: Influenza Vaccination Coverage, Centers for Disease Control and Prevention. <https://www.cdc.gov/flu/fluvaxview/interactive-general-population.htm>. Accessed on 24 August, 2023.
